# Sudan’s complex genetic admixture history drives adaptation to malaria in Sudanese Copts

**DOI:** 10.1101/2025.09.01.25334828

**Authors:** Laura Vilà-Valls, Jorge Garcia-Calleja, Javier Prado-Martínez, Elena Bosch, Aida M. Andrés, Mihai G. Netea, David Comas, Hisham Y. Hassan

## Abstract

Sudan lies at the crossroads of Africa and the Middle East, with rich cultural, linguistic, and ecological diversity shaped by a complex demographic history. We present the first whole - genome sequencing (WGS) study of Sudanese populations, analyzing high-coverage genomes (∼30×) from 125 individuals representing five ethnolinguistic groups across three language families. Our results reveal deep population structure, involving Nilo-Saharan, West Eurasian, Northern African, and Western African ancestral components, as well as signatures of the Arab expansion. We report over one million novel variants, including population-specific deleterious alleles, highlighting the need for broader African genomic representation. Notably, local ancestry inference reveals a strong signal of adaptive admixture on chromosome 1 in Sudanese Copts, marked by a peak of Nilo-Saharan ancestry introduced via genetic admixture 1,000-1,500 years ago. At this locus, we estimate a remarkably strong selection coefficient (s=0.0996) for SNP rs2814778 within the *ACKR1* gene, which is responsible for the Duffy-null blood group that provides resistance to *Plasmodium vivax* malaria. These findings reveal Sudan as a genomic mosaic shaped by ancient and recent migrations and provide clear evidence of admixture-driven adaptation in an understudied region of Africa.

**Significance:** Despite their rich cultural, linguistic, and genetic diversity, Sudanese populations have been largely underrepresented in human genetic research, particularly in whole-genome sequencing (WGS) datasets. Here, we present 125 new high-coverage genomes from Sudan and uncover a complex history of admixture shaped by major human migrations, including the Arab expansion and the spread of Nilo-Saharan speakers. We also find strong evidence of post-admixture adaptation to malaria, and report over one million novel genetic variants. This study not only fills important gaps in our understanding of Africa’s demographic history but also highlights the importance of including recently admixed populations to better understand human evolution and local adaptation.

## Introduction

Located in Northeast Africa and bordering seven countries and the Red Sea, Sudan is the third largest country on the African continent, home to over 40 million people. Its predominantly arid climate, ranging from desert in the north to semi-humid zones in the south, is characterized by limited and irregular rainfall and temperatures often exceeding 40°C. These harsh conditions are challenging for its inhabitants, who primarily depend on crop and livestock farming. The Nile River, the world’s longest, serves as Sudan’s primary water source. It supports agriculture and human settlement in an otherwise water-scarce landscape(1). Sudan’s strategic position has made it a cradle of civilizations, with the Nile functioning as a corridor for trade and demographic movements, linking Egypt and the Mediterranean Sea with Eastern Africa(2). The oldest modern human remains found in Sudan (Singa) date to the Late Middle Pleistocene (∼133 ka)(3), while archaeological evidence suggests over 300,000 years of human presence in the region(4, 5).

As a result of successive demographic movements, Sudan is currently home to approximately 600 ethnic groups, speaking over 400 languages and dialects. These include three of Africa’s major language phyla: Afro-Asiatic, Niger-Congo and Nilo-Saharan, as well as the Kordofanian putative language isolates of southern Sudan. Despite this linguistic diversity, Arabic is the official and most widely spoken language(1). Afro-Asiatic languages dominate northern and eastern Africa. In Sudan, this phylum includes Arabic and Cushitic languages, primarily found in the Horn of Africa, such as Beja, spoken by nomadic communities in northeastern Sudan. Beja people are organized into tribes and mainly inhabit the mountainous range between the Red Sea and the Nile and Atbara rivers, in the Eastern Desert. They are primarily pastoralists, and their herds of cattle and camels are essential for their subsistence(6, 7). Coptic also belongs to the Afro-Asiatic language family and is the native language of Copts, the largest Christian population in the Middle East and North Africa, who primarily reside in Egypt, Sudan, and Libya(6, 8, 9). The major introduction of Christianity into Egypt and Nubia occurred during the 6th century CE(10, 11). Coptic derives from Ancient Egyptian and is now used almost exclusively in liturgical contexts, as the Coptic population was substantially influenced by Arabization(6, 8, 9). After the Arab conquest of Egypt in the 7th century, Coptic Christians who did not convert to Islam faced persecution, leading some to migrate beyond Egypt(9). Nilo-Saharan languages are spoken across Eastern and Central Africa. In Sudan, they are spoken by Nubian people, such as Mahas, and by inhabitants of Darfur, in western Sudan. Nubians are divided into different ethnolinguistic groups, who historically lived along the Nile River in southern Egypt and northern Sudan, although many of them moved to urbanized areas and adopted Arabic and Islam(1, 6, 12). Darfur meaning “the home of the Fur”, takes its name from Fur, the region’s indigenous people. This ethnolinguistic group lives in the highest region of the country, the Jebel Marra massif. Historically, they established powerful kingdoms until the

Arab arrivals pressure pushed them to more isolated places, which also preserved their identity and culture. They are traditionally agriculturalists, and cultivate a variety of cereals and crops. After some nomadic Arab pastoralists moved to Darfur, discrepancies over land and water access started tensions that escalated into war conflict in 2003, which continues today(6, 13, 14). Niger-Congo languages are represented by Fulfulde, spoken by Fulani, nomadic pastoralist groups with a widespread presence across the Sahel, from the Atlantic Ocean to western Sudan, and strong roots in Western Africa. Traditionally they herd cattle, goats and sheep, mostly for dairy production. In recent times, some groups have adopted a semi-sedentary or fully sedentary lifestyle. Historical records indicate that they started to move from Futa Toro (Senegal) in the 10th century CE, reaching Hausaland around the 15th century, and Darfur in the 18th century. As they migrated, the Fulani interacted with Arab culture, and some became missionaries. The population is estimated at about 25 million, with communities dispersed in smaller groups across some regions. Despite this, intermarriage remains widely accepted among different Fulani groups(1, 6, 15).

Genetic studies have revealed that African populations exhibit extensive genetic diversity, structured into multiple ancestries; defined as lineages tracing back to genetically differentiated ancestral populations. These ancestries often correlate with ethnic, linguistic, and cultural groups. These studies also reflect complex histories of migration and gene flow, as most African populations show substantial admixture(16, 17). In northeastern Africa, mitochondrial DNA evidence from early research pointed to the Nile River Valley as a key driver of such demographic events, including long-term, bidirectional gene flow between North Africa and the rest of the continent(2). In Sudan specifically, genetic studies have identified multiple ancestral components, including autochthonous ancestries in Nilo-Saharan speakers and Fulani pastoralists(18–22). Recent work places the origin of Fulani in the ‘Green Sahara’ during the most recent African Humid Period (12,000–5,000 BP)(23, 24). During these recurrent humid phases, the Sahara Desert became a green savanna with vegetated corridors and connected lakes and rivers, that facilitated fauna and hominin dispersals. The last of these episodes favored the spread of pastoralism in Africa, and the domestication of cereals such as sorghum, first domesticated in eastern Sudan 4,000 BCE(25–28).

A significant contribution of Middle Eastern-like ancestry has also been identified in Sudan(18, 19, 22), likely introduced during the Arab expansion beginning in 639 CE. This genetic signal is particularly strong among groups such as Beja and Nubians, in line with genealogical and cultural records(1, 6). Additionally, a Western African component entered Sudan during the past 300 years(19, 29, 30).

Ancient DNA from a Christian-period Nubian cemetery (∼650–1,000 CE) in present-day Sudan provides direct evidence of historical gene flow between Egypt and Nubia(21). Individuals from this period exhibited comparable proportions of Nilo-Saharan and West Eurasian ancestries, with the latter likely introduced through Egypt. In contrast, this Nilo-Saharan component is absent in Ancient Egyptian samples(31) and remains limited in modern Coptic Christians, consistent with its Egyptian origin(19, 22).

These findings suggest that gene flow between Egypt and Nubia predates the Arab expansion, occurring as early as 111-265 CE(21). Present-day Nubians, in contrast, show more recent admixture likely reflecting interactions during the Arab expansion(19, 21).

Despite the region’s challenging arid environment, northeastern Africa has supported continuous human habitation for millennia. Both autochthonous and incoming populations have likely been exposed to a range of selective pressures that may have driven genetic adaptations to local environmental conditions, such as endemic disease, UV exposure and diet. Although some adaptive signals have been reported, the role of selection in this region remains largely underexplored. Notably, pastoralist groups such as Fulani and Beja harbor multiple lactase persistence (LP) mutations associated with dairy consumption(32, 33). In Sudan, additional selection signatures have been identified in genes related to immune responses against malaria and bacterial infections(22). Due to its ecological diversity and relatively greater water availability compared to adjacent Saharan zones, parts of Sudan offer favorable conditions for mosquito proliferation, leading to higher exposure to vector-borne diseases like malaria(34, 35). The ACKR1 gene, associated with *Plasmodium vivax* malaria resistance, was found to be enriched of African ancestry for admixed Arab and Nubian populations from the Eastern Sahel(36). Additionally, Fulani show distinct immune responses against the most severe forms of malaria, showing fewer symptoms than neighbouring populations, suggesting population-specific genetic adaptations(15, 37). In Ethiopian populations, signals of adaptation have also been found in genes related to folate metabolism, skin pigmentation, and UV protection(38).

In this study, we analyzed 125 newly sequenced high coverage genomes from five Sudanese populations spanning three language families. Our findings provide evidence of post-admixture adaptation to malaria in Sudanese Copts, resulting from admixture between Middle Eastern and Eastern African populations approximately 1,000-1,500 years ago. These results offer a valuable foundation for future research into the mechanisms of rapid human adaptation in this understudied region.

## Results

### Whole-genome variation in Sudan

We generated to average sequencing coverage of ∼30X whole-genome sequencing (WGS) data for 125 individuals from five Sudanese populations: Beja (N=25), Copts (N=25), Fulani (N=25), Fur (N=25) and Mahas (N=25). These groups represent three distinct language families: Afro-Asiatic, Nilo-Saharan and Niger-Congo (*SI Appendix*, Dataset S1). After rigorous quality control, including filters for sequencing depth, genotype quality, and missingness, we retained 24,774,084 biallelic SNPs.

Of these, 1,136,610 are novel SNPs not reported in dbSNPv156(39), gnomADv4.1(40), or the HGDP+1KG callset(41). As expected, the majority of these previously unreported SNPs are singletons (77.4%) and population-specific (86.2%), with Fur population harboring the highest number of novel population-specific variants.

Additionally, we detected 1,542 novel predicted deleterious variants, including 1,352 amino acid substitutions predicted to be probably or possibly damaging by PolyPhen-2(42), 102 stop codon gains/losses, 7 start codon losses, 80 splice acceptors/donors and a downstream gene variant classified as likely pathogenic in ClinVar(43). Of these, only 14 have a derived allele frequency greater than 0.05 (*SI Appendix*, Table S1).

### Population substructure

We analyzed this high-coverage WGS dataset, together with a broad reference panel (*SI Appendix*, Dataset S2), to investigate the population structure of Sudan. Principal Component Analysis (PCA) confirms deep genetic structure within Sudan. Fur individuals show genetic affinities to other Eastern African populations, whereas Beja and Mahas form distinct yet proximate groups that fall between Eastern Africans and West Eurasians, suggesting potential admixture. Copts fall tightly with Middle Eastern populations, while Sudanese Fulani form a cline with other Fulani groups spanning from Western to Northern and Eastern Africa, with no clear separation by country (Fig. 1 *A*).

**Fig. 1.**
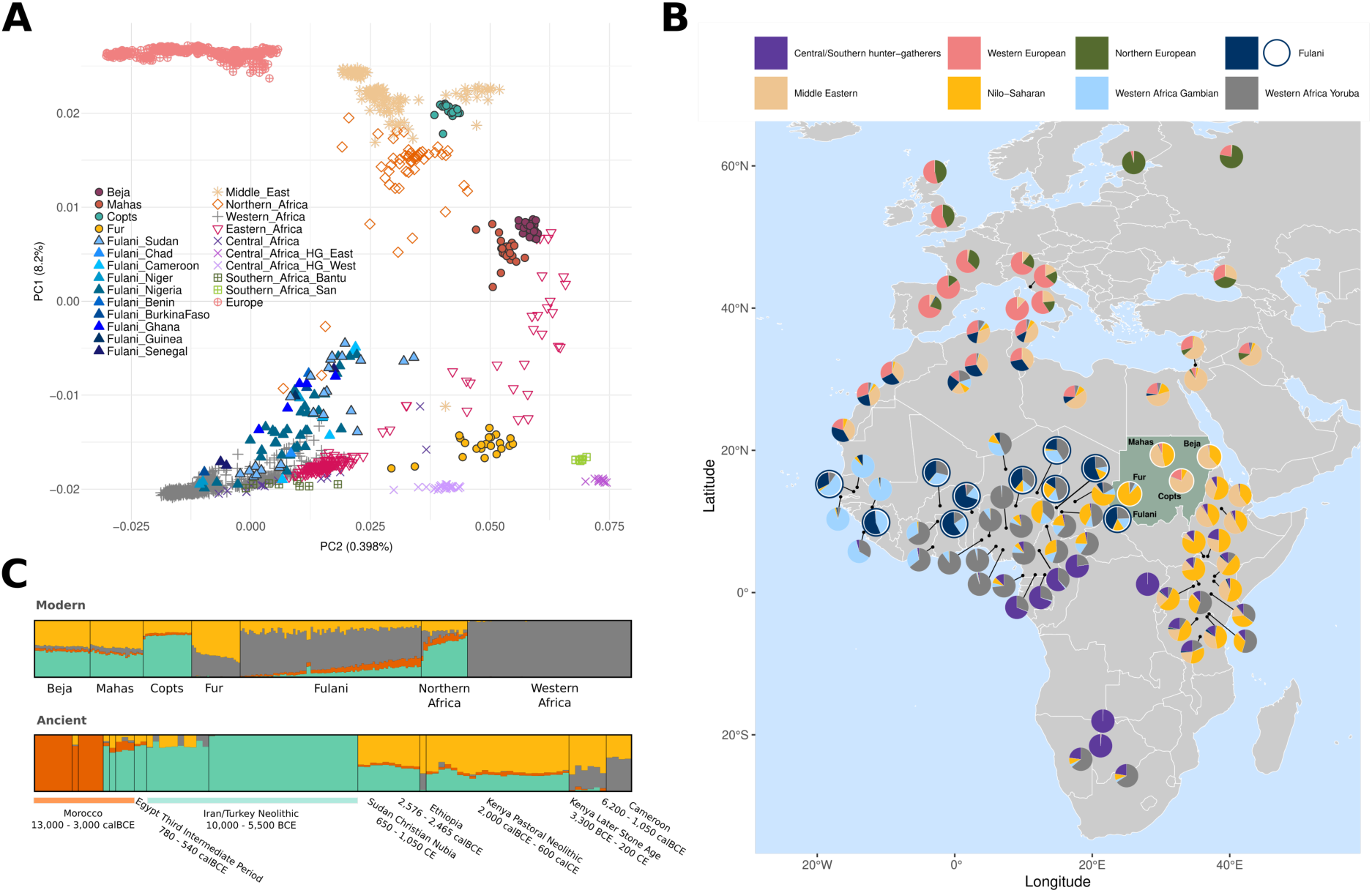
Genetic structure of Sudan with PCA and ADMIXTURE. (*A*) PCA for modern WGS data, with PC1 against PC2. Sudanese and Fulani samples are represented with filled shapes. Some populations are grouped for simplification. Central_Africa_HG stands for Central African rainforest hunter-gatherers, grouped as in Fan et al. (2019)(78). (*B*) ADMIXTURE map for WGS data, for the lowest cross-validation error (K=8). Populations located at the same coordinates are grouped together. For populations where only the locality or country is known, the coordinates are approximate. (*C*) ADMIXTURE analysis with ancient DNA, showing the first four components (K=4). The lowest cross-validation error was for K=2. At the top, modern populations using as Western Africa reference Esan from Nigeria. At the bottom, ancient populations. Ancient Moroccan samples are ordered from left to right as follows: Epipalaeolithic from Taforalt (TAF), Epipalaeolithic from Ifri Ouberrid (OUB), Early Neolithic from Ifri n’Amr o’Moussa (IAM), Early Neolithic from Kaf Taht el-Ghar (KTG), Middle Neolithic from Skhirat-Rouazi (SKH) and Late Neolithic from Kehf el Baroud (KEB).

To integrate the important linguistic component to this analysis, we incorporated genotype array data, mainly from Eastern Africa, spanning different language families into a broader PCA analysis. While Fur and other Nilo-Saharan speakers form a genetic cline, not all populations group according to their language family. Notably, Mahas, despite being Nilo - Saharan speakers, appear close to Sudanese Afro-Asiatic speakers (*SI Appendix*, Fig. S1). Unsupervised ADMIXTURE analysis(44) identified six distinct genetic components in Sudanese populations (Fig. 1 *B*). A predominant Nilo-Saharan-like component, widespread in Central and Eastern Africa, is most frequent in Fur (86.5%) and present at varying levels in Beja, Mahas, Copts, and Fulani. A Middle Eastern/North African component is most prevalent in Copts. This component is also present in Beja and Mahas. Fulani show a third, distinct component also found in Northern Africa. Additional ancestries include a European-like component in Copts and Western African components in Fulani and Fur (Fig. 1 *B*). Ancient DNA data reveals Northern African components related to Epipalaeolithic Iberomaurusians and Neolithic back-to-Africa movements, in Beja, Mahas, Copts, and Fulani, along with a widespread Nilo-Saharan-like ancestry in ancient Eastern and Central African individuals (Fig. 1 *C*), highlighting Sudan’s complex admixture history shaped by both ancient and recent admixture events.

Estimates of effective population size (N_e_) trends (*SI Appendix*, Fig. S2) and runs of homozygosity (ROH) (*SI Appendix*, Fig. S3) reflect increased genetic drift in Copts, likely driven by cultural and/or geographic isolation compared to the other Sudanese groups. The prevalence of short ROH segments suggests past inbreeding, a pattern also observed in West Eurasian populations.

We further examined the genetic substructure within Sudan using haplotype-based approaches, including ChromoPainterv2(45) and fineSTRUCTURE(45). Sudanese individuals display considerable genetic substructure, generally forming population-specific branches in the resulting dendrogram (*SI Appendix*, Fig. S4). Fulani and Fur individuals cluster with the majority of African populations, whereas Beja, Copts and Mahas group within a separate branch predominantly composed of West Eurasian populations, along with North Africans. Notably, Fulani form two distinct branches: *Fulani_East*, comprising primarily Fulani and two Nigerian Kanuri individuals, and *Fulani_West*, clustering with other Western and Central African populations (*SI Appendix*, Fig. S4 and Dataset S3).

### Sources of admixture in Eastern Africa

To infer the timing of admixture events, we first grouped individuals into genetically homogeneous clusters based on fineSTRUCTURE analysis (*SI Appendix*, Fig. S4 and Dataset S3). The average total variation distance (TVD) between clusters was 1.296, with a minimum of 0.007 (*SI Appendix*, Dataset S4). PCA on the ChromoPainter coancestry matrix confirmed that haplotype-based clusters capture underlying genetic structure more effectively than population or linguistic labels, which often group genetically diverse individuals together (*SI Appendix*, Fig. S5). The haplotype-based PCA revealed clustering patterns similar to those observed in the allele frequency-based PCA (Fig. 1 *A*), highlighting pronounced genetic structure within East Africa.

We then applied fastGLOBETROTTER(46) and MALDER(47), using Fur as a representative Nilo-Saharan donor population. With both methods, we detected overlapping admixture dates for all recipient clusters, except for *Fulani_West*, although the difference is less than 500 years (∼15 generations) (Fig. 2 and *SI Appendix*, Table S2) . In both Beja and Mahas, we identified Northern African and Middle Eastern genetic components, using SOURCEFINDv2(48) (*SI Appendix*, Fig. S6), and detected Arabic-related admixture dating to 1,512 years ago in Mahas and 840 years ago in Beja, based on admixture events estimated at ∼54 and ∼30 generations ago, respectively (assuming 28 years per generation; (Fig. 2 and *SI Appendix*, Table S2). These admixture events involved a major Eastern African source, comprising Nilo-Saharan and other Eastern African clusters, and a minor source from North African/Middle Eastern populations. Mahas show a higher proportion of Nilo-Saharan ancestry, whereas Beja exhibit more Eastern African ancestry, supported by their different linguistic and historical backgrounds (*SI Appendix*, Dataset S1).

**Fig. 2.**
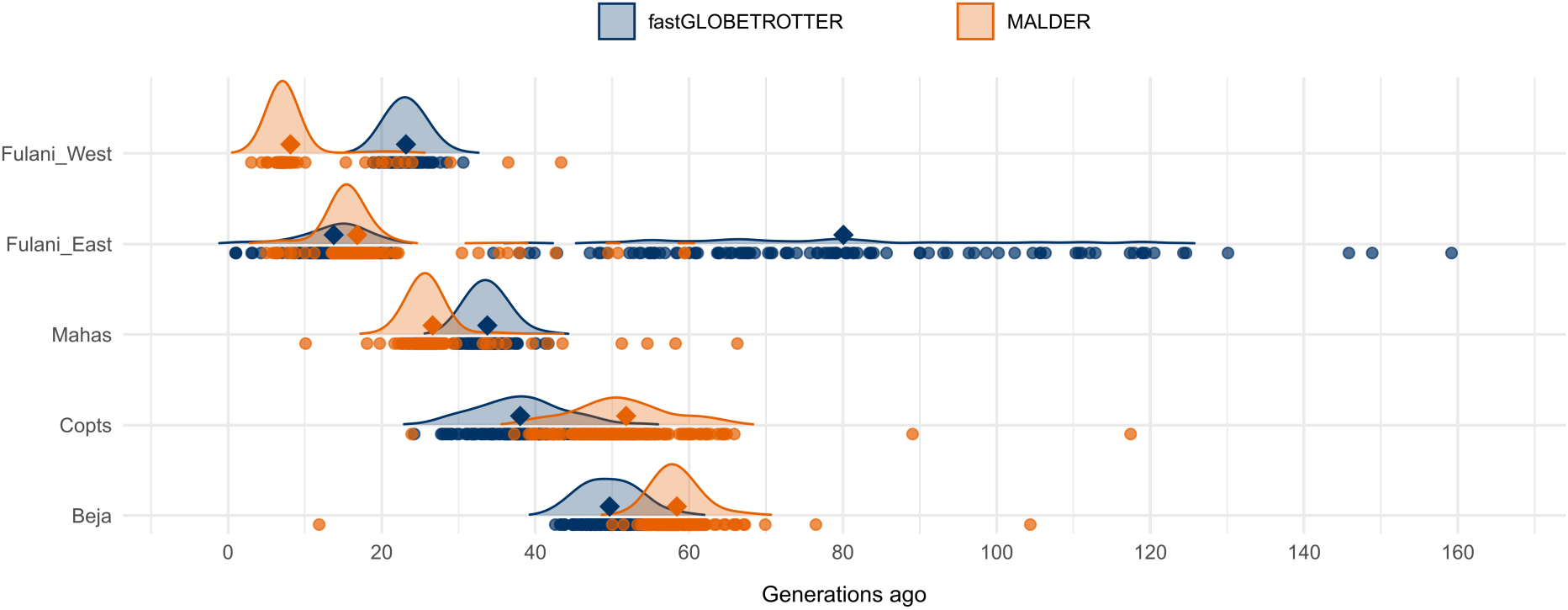
Distributions of admixture dates inferred using fastGLOBETROTTER and MALDER. Admixture dates were estimated for the target genetic clusters defined with fineSTRUCTURE using two different methods. For fastGLOBETROTTER, 100 bootstrap estimates per admixture event are shown as dots, while for MALDER, dots represent significant admixture events detected between pairs of source populations. Diamonds indicate the average admixture date for each method. For *Fulani_East* cluster, multiple waves of admixture were inferred with fastGLOBETROTTER. Time is represented by generations before present.

Copts show predominantly Middle Eastern and Northern African ancestry, with smaller (<2%) Nilo-Saharan and Southern European components (*SI Appendix*, Fig. S6). Slight Nilo-Saharan-like contribution (*SI Appendix*, Fig. S6) dates to ∼45 generations ago (37-53 GA), while MALDER identified two older admixture events (89 and 117 GA, or ∼468 and 1,252 BCE), involving Northern African and European sources (Fig. 2 and *SI Appendix*, Table S2). Both Fulani genetic clusters (*Fulani_East* and *Fulani_West*) have a major contribution from Western African sources, with *Fulani_East* carrying more Northern/Eastern African ancestry compared to *Fulani_West* (*SI Appendix*, Fig. S6). The dates of admixture suggest a single event for *Fulani_West* around 16 GA and multiple waves for *Fulani_East* around 15 GA and 80 GA (Fig. 2 and *SI Appendix*, Table S2). However, it can be interpreted as continuous admixture, since the source populations show similar ancestry proportions (*SI Appendix*, Fig. S6).

Using modern populations as donor groups, our analyses indicate admixture in the last two millennia among Fulani populations across the Sahel, including Sudan. However, previous studies(23, 24) suggest that some of these admixture events may trace back to the Green Sahara period. To address this question with our new Fulani samples from Sudan, we used DATES(49). Ancient DNA from Cameroon (∼7,000 BP)(50) was used as a Western African source, Moroccan Early Neolithic (IAM) individuals (∼6,000 BP)(51) as a Northern African source, and Later Stone Age individuals from Kenya (∼4,000 BP)(52, 53) as an Eastern African source, following previous approaches(23, 24). Admixture between ancient Western Africans and ancient Northern Africans is inferred at 266±180 GA (2,511-12,935 ya) and between ancient Western Africans and ancient Eastern Africans at 228±153 GA (2,173-11,045 ya), supported by an exponential decay of ancestry covariance (*SI Appendix*, Fig. S7), and suggesting additional admixture events in Fulani predating the dates inferred using modern genomes.

### Candidates for positive selection in admixed populations

Sudan’s strategic position and historical role as a corridor for human migrations make its admixed populations relevant for investigating adaptation to local environments. While much of Sudan is arid and semi-arid, the presence of major rivers and transitional zones between desert and grassland savanna(1) introduces ecological variability that may have led to very local selective pressures, including exposure to endemic pathogens and other environmental challenges.

Because Sudanese groups display complex admixture patterns involving West Eurasian, Nilo-Saharan, and Western African ancestries, investigating the distribution of these ancestral components across the genome allows us to test whether ancestry-specific genomic segments were favored by natural selection following admixture. We applied RFMix(54) to estimate the local distribution of West Eurasian, Nilo-Saharan and Western African ancestries, and identify genomic regions where ancestry components are significantly over- or underrepresented relative to the population average (referred to as *local ancestry deviations,* LADs). These deviations may reflect post-admixture adaptive processes, where certain alleles inherited from a specific ancestral population may have conferred a selective advantage in the post-admixture population.

The most significant LAD across all populations, and the highest z-score after discarding peaks at chromosome ends that are prone to technical artifacts, was found in Copts on chromosome 1 q23.1-q23.2, spanning ∼2.2 Mbp (*SI Appendix*, Dataset S5). While the genome-wide average of Nilo-Saharan ancestry in Copts, based on RFMix local ancestry inference, is only 4.3%, this locus shows a marked increase to 29.7%, a seven-fold increase that corresponds to 6.9 standard deviations above the mean. This is accompanied by a reciprocal decrease in West Eurasian ancestry (z-score = -6.3), and a slighter increase in Western African ancestry (z-score = 2.64) (Fig. 3 *A* and *SI Appendix*, Fig. S8). This genomic region includes the SNP rs2814778 (T>C) (chr1:159204893, GRCh38), located in the promoter region of the *ACKR1* (atypical chemokine receptor 1) gene. The derived (C) allele is responsible for the Duffy-null blood group, which leads to the absence or reduction of the Duffy antigen expression on the surface of red blood cells. This absence prevents *Plasmodium vivax*, a malaria-causing protozoan, from binding and entering erythrocytes, thereby conferring resistance to infection. The rs2814778 (C) or Duffy-null allele represents one of the strongest known signals of recent positive selection in humans, with striking geographic variation, nearly fixed across the African continent but rare in populations elsewhere(55). Although no population-level data were previously available for the Sudanese groups analyzed here, selection at this locus has been shown in other admixed populations(56–59), supporting Duffy-null as the most plausible driver of selection in this region.

**Fig. 3.**
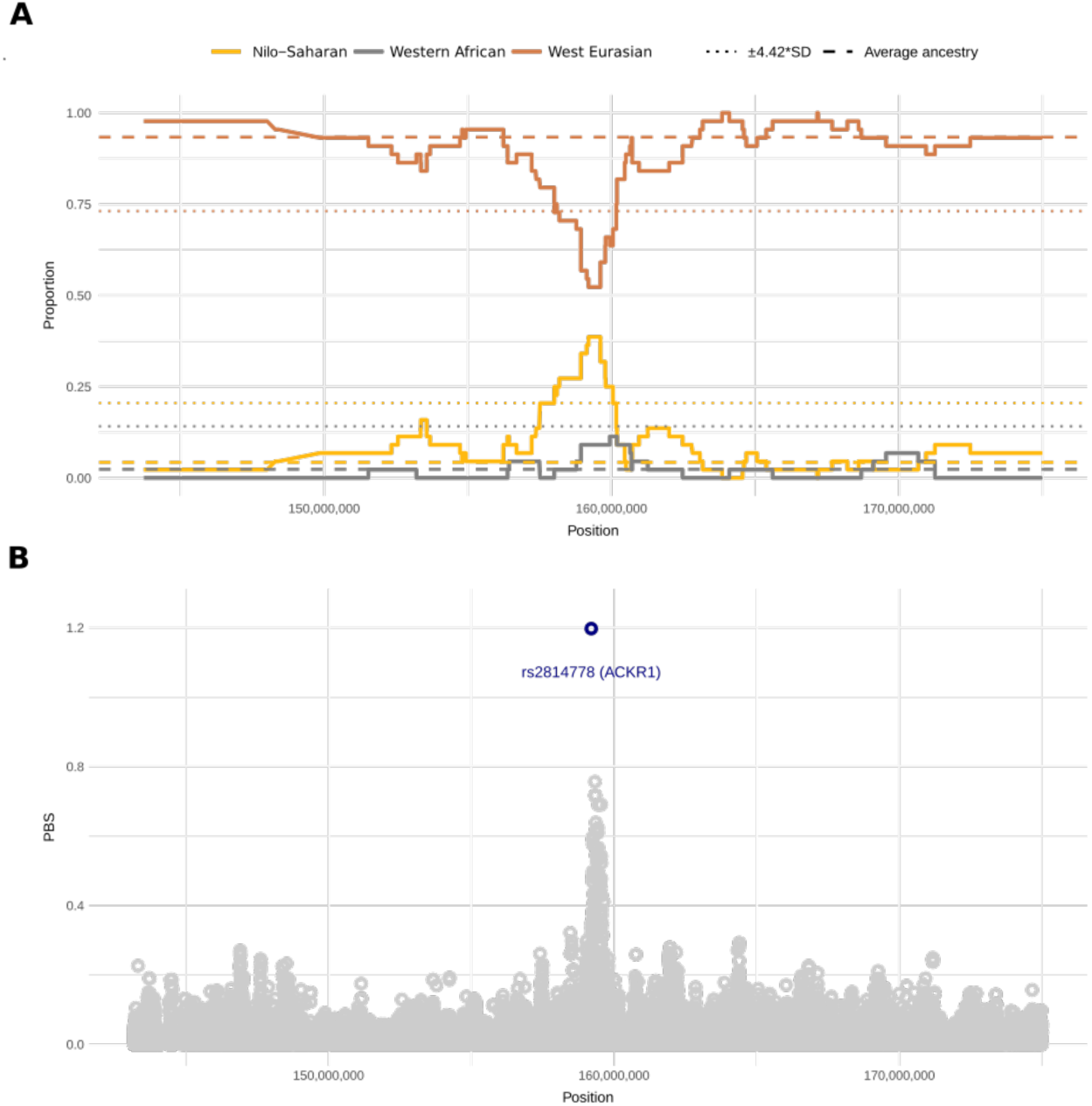
Selection signal on Copts chromosome 1. (*A*) region of chromosome 1 comprising the highest genome-wide Local Ancestry Deviation (LAD) values: Nilo-Saharan, represented by Fur; West Eurasian, represented by CEU and Bedouin; and Western African, represented by Yoruba. (*B*) Population Branch Statistics (PBS) estimates for the same genome region. In blue the SNP and its gene with the highest PBS genome value for Copts.

Nevertheless, the extent of the selected region (∼2.2 Mbp) and the presence of over 100 variants classified as deleterious with VEP(84), suggest that other loci may also have been involved in, or impacted by, this selection process. Functional annotation revealed enrichment of immune-related GO terms, within others (*SI Appendix*, Dataset S5). Additionally, alleles particular to African ancestry have previously been associated with cardiovascular disease risk, asthma susceptibility and low white cell counts(56, 60–62). Notably, some of these alleles are in linkage disequilibrium with the Duffy-null allele, suggesting that positive selection on Duffy-null may have influenced other relevant loci, and although Dufffy-null appears to be the primary target, this region could have a broader immunological and health-related consequences(56).

We tested whether this region had LAD in Beja and Mahas using a lower z-score threshold of ±2. It was significant in Mahas, with z-scores of 2.51 and -3.07 for the Nilo-Saharan and West Eurasian ancestries, respectively (*SI Appendix*, Fig. S8). Under this more relaxed threshold, Fulani also showed a significant increase (z-score of 2.02) in West Eurasian ancestry on chromosome 2, spanning the *LCT* and *MCM6* genes, consistent with the frequency of the Eurasian lactase persistence (LP) variant 13910*T previously reported in Fulani. In contrast, Beja did not show ancestry deviation in this region, likely because lactase persistence in this group is driven by variants of different origin, such as 14009*G, found in Ethiopia, which aligns with their Cushitic origin(32, 33, 63).

Additional LADs were identified in Copts, Beja and Fulani (*SI Appendix*, Dataset S5). Most of these regions are associated with functions related to chromatin and DNA organization, gene expression, immune response, and transport activity.

SNP rs2814778 in Copts appears as the strongest candidate for positive selection across the genome when analyzing Beja, Coptic, and Mahas populations using Population Branch Statistics (PBS)(64) (Fig. 3 *B* and *SI Appendix*, Dataset S6). Additional peaks encompass genes involved in chromatin and transcription regulation, membrane activity, transport, immune response, alcohol metabolism, and structural integrity (*SI Appendix*, Fig. S9 and Dataset S6).

We further explored the rs2814778 (C) allele frequency across the region (Fig. 4 *A*). On average, this has a frequency of 92% in Sudan. The derived allele is fixed in both Fulani and Fur individuals, and its frequency exceeds 0.8 in Beja, Copts, and Mahas. Notably, Copts exhibit a frequency of 0.89, substantially higher than that observed in West Eurasian populations (e.g. Bedouin 0.34, Druze 0.01, IBS 0.01, GBR 0).

**Fig. 4.**
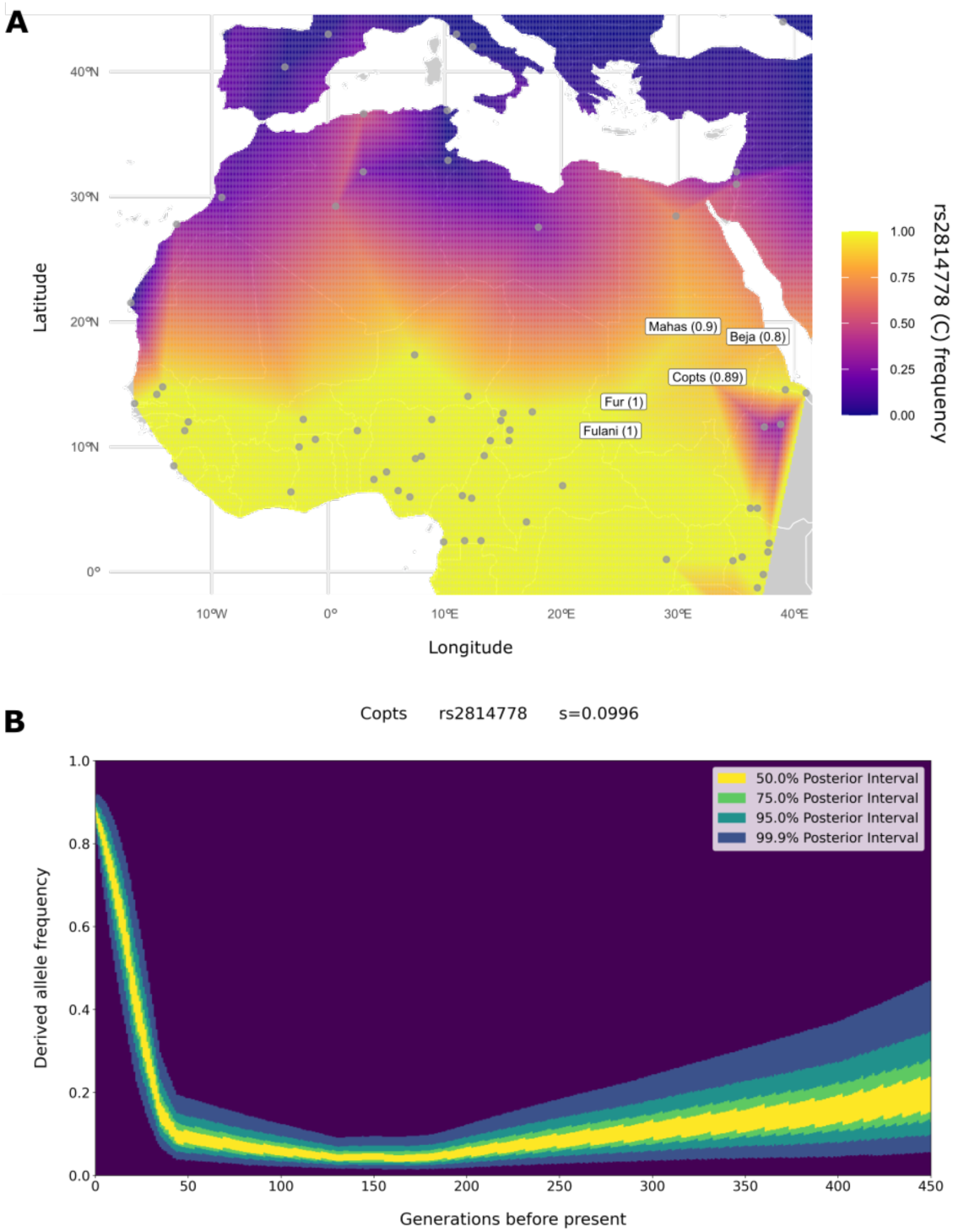
Allele frequency and selection of the Duffy-null allele. (*A*) interpolation map for the allele frequency of rs2814778 (C) or Duffy-null allele. Gray dots represent the populations used in their coordinates, the same as in Fig. 1. For populations with common coordinates, we calculated the aggregate allele frequencies. Labels correspond to Sudanese populations and their allele frequencies. (*B*) Derived allele frequency trajectory and selection coefficient (s) of rs2814778 for Copts, computed with CLUES2 for the epoch 0-45 GA.

The allele frequency trajectory of rs2814778 (C) over time and the selection coefficient were computed with CLUES2(65, 66). Selection was inferred between the present and 45 generations ago, corresponding to the average admixture time estimated using fastGLOBETROTTER and MALDER. The selection coefficient of rs2814778 (C) estimated for Copts is 0.0996 (Fig. 4 *B*), with 95% confidence within 0.09001-0.10920 and a logLR of 19.3115, suggesting remarkably strong post-admixture selection occurring within a timespan of less than 1,500 years. With a classical formula from Hartl & Clark (1997)(67), based on allele frequency change over time and assuming an additive fitness effect of the Duffy-null allele(55, 68, 69), the selection coefficient was estimated to be 0.208. For this calculation, we used an initial rs2814778 (C) frequency of 0.07, derived from admixture proportions in the modern Coptic population from Sudan and the source contributions estimated at the time of admixture with fastGLOBETROTTER, and applied the same timescale as in CLUES2 (45 generations).

## Discussion

In this study, we generated and analyzed high coverage whole-genome sequencing (WGS) data from 125 individuals representing five Sudanese populations across three language families. We identified approximately 1.1 million previously unreported variants, of which 1,542 are predicted to be damaging, highlighting the need for broader representation of African populations in human genomic research.

We observed substantial genetic structure within Sudan, in line with earlier findings(18, 19, 22), but with a clearer differentiation between populations than previously reported. Distinct genetic components were identified for Nilo-Saharan speakers from Darfur and for Fulani pastoralists, consistent with previous studies(18–22). Nilo-Saharan ancestry appears broadly distributed across Eastern and Central Africa, reflecting the historical range of Nilo-Saharan speakers. Its presence in ancient individuals supports deep origin, likely linked to the spread of Nilo-Saharan languages (∼8,000 BCE)(70).

Fulani show a distinct genetic component(22–24), also found in Northern Africa, reflecting admixture between Northern and Western African groups. This is supported by the detection of a genetic signal similar to that of Epipalaeolithic Iberomaurusians (∼13,000 BP, Morocco), reinforcing their Northern African ancestry. Fulani also carry a Nilo-Saharan component, more evident in those from the eastern Sahel. Our analysis indicates recent gene flow from Northern/Eastern African sources around 400 years ago. Additionally, we identified older admixture during the last African Humid Period, aligning with previous studies(23, 24), which suggests an ancestral connection to populations linked to the spread of pastoralism during favorable climate conditions in the Sahara Desert, further supported by ancient genomic data (∼7,000 years old) from Takarkori, Libya(28). The Green Sahara not only facilitated the spread of pastoralism but also provided the ecological setting for the dispersal of Nilo-Saharan-speaking groups, who initially relied on aquatic resources and food collecting but later transitioned toward farming(26, 70). Therefore, this period likely enabled interactions among at least three ancestral groups: Western Africans, Northern Africans, and ancient Nilo-Saharan populations.

Copts, a Christian ethnoreligious group with origins in Egypt, primarily exhibit West Eurasian ancestry, along with distinctive Eastern African gene flow. This admixture, dated to 1,000-1,500 years ago, may be associated with the spread of Christianity into Nubia, beginning around 542 CE(10, 11). Genetic evidence of Egypt-Nubia contact during the early Christian Period (∼650-1,000 CE)(21) overlaps with our admixture estimates. We also identified two earlier admixture events involving Northern African and European sources, suggesting historical Mediterranean connections. Additionally, evidence of past inbreeding among Copts, reflected in ROH patterns and reduced effective population size, is consistent with a founder effect following their migration into present-day Sudan.

Nubian Mahas, a Nilo-Saharan speaking group, and Beja, Afro-Asiatic speakers from the Eastern Desert, show greater genetic affinity with Eastern African Afro-Asiatic populations. Despite their linguistic divergence, both groups are genetically similar and appear to be admixed between a Northeast African and a West Eurasian population. Although allele frequency-based analyses did not reveal major genetic differences between Mahas and Beja, haplotype-based methods showed that Mahas harbor higher Nilo-Saharan ancestry, whereas Beja display greater affinity to Eastern Africans from the Horn of Africa. This pattern is consistent with their respective Nubian and Cushitic background. The West Eurasian genetic component has been linked to the Arab expansion following the arrival of Islam in Egypt in 639 CE(22), though an earlier contribution associated with the introduction of Christianity in Nubia is also possible(10, 11). Mahas show a more recent inferred admixture date (∼840 years ago) compared to Beja (∼1,512 years ago). These dates are comparable to previous observations of Arabic-related admixture in other Nubian and Cushitic groups(18, 71). The populations’ Islamic faith and the historical influence of the Arab migration suggest that their non-African ancestry primarily came from the Middle East. However, the presence of an ancient Northern African genetic component indicates that the West Eurasian source had contact with Northern Africans before reaching Sudan, aligning with the timeline of the Arab expansion in Africa(6). Nevertheless, this result should be interpreted with caution due to the technical limitations associated with ancient DNA analyses. While some studies have proposed that Beja’s contact with West Eurasian populations occurred earlier via sea routes through the Arabian Peninsula(19), in our analysis we only detect one estimate older than 100 GA.

While these admixture events have shaped the genetic composition of the studied populations, they may also have introduced alleles that, under certain selective pressures, such as environmental or pathogenic challenges, contributed to local adaptation. In the absence of selective pressures, ancestral components are generally expected to be uniformly distributed throughout the genome. However, Copts exhibit a pronounced excess of Nilo-Saharan ancestry on chromosome 1, suggesting that Nilo-Saharan alleles were strongly favored by positive selection post-admixture. This is further supported by pronounced genetic differentiation from the nearest population, Bedouin Arabs, at the top candidate SNP rs2814778 (T>C) in the *ACKR1* gene (FST = 0.474, compared to a genome-wide FST = 0.011), along with a strong selection coefficient (0.0996). This variant is of functional and phenotypic importance that is linked to fitness because it results in the Duffy-null allele that confers resistance to *Plasmodium vivax* malaria and is widely recognized as a hallmark of recent positive selection in other human populations(55–59). The Duffy-null allele is the most plausible driver of this selection signal, yet additional factors may have contributed to the marked increase of Nilo-Saharan ancestry at 1q23(56, 60–62). In Mahas, Nilo-Saharan ancestry was also selected in this region considering a lower threshold, in accordance with previous observations in other admixed Nubian groups(36).

We report for the first time the frequency of the derived allele (C) in Sudanese populations, averaging 92% across Sudan, being fixed in two populations with different demographic histories, Fur and Fulani, and 89% in Copts, of Egyptian origin. The latter is consistent with the observed excess of Nilo-Saharan ancestry and the strong genetic differentiation at the Duffy-null locus. Given these findings, along with the near fixation of the Duffy-null allele across the African continent south of the Sahara Desert and its rarity in West Eurasia(55), the most plausible explanation is recent positive selection acting on the allele after admixture. Similar cases of post-admixture selection on the Duffy-null variant have been documented in other populations, including Pakistan(58), Madagascar (with selection coefficients of s=0.066(59) and s>0.2(56)), and Cabo Verde (s=0.08)(57).

Taken together, these findings align with a scenario where, following admixture with an Eastern African population less than 1,500 years ago, swift selective forces favored the retention and subsequent increase of Nilo-Saharan-like ancestry at this locus in Copts, conferring an adaptive advantage against malaria. This constitutes one of the strongest documented signals of selection on the Duffy-null allele.

The prevalence of *Plasmodium vivax* in Eastern Africa today could explain the rapid post-admixture adaptive selection observed in Copts. While the pathogen is almost absent across much of the African continent due to the high frequency of the Duffy-null allele and the limiting conditions for vector survival in the north, Sudan and Ethiopia reported more than a million cases in 2017(72). Ancient DNA provides additional context: *P. falciparum* has been identified in human remains from Ancient Egypt dated to 1,500-500 BCE, and a single case of *P. vivax was recovered* from 800-573 BCE (73–75). Phylogenetic evidence indicates that *Plasmodium vivax* originated in African great apes(76), while the most common Duffy-null haplotype has been estimated to be ∼42,000 years old(55). These findings, although limited by aDNA preservation, raise important questions about the history of malaria in the region, and whether selective pressures arose in the first civilizations of Northeast Africa, or are the response of more recent exposures to the pathogen. Moreover, without comparable genetic data from modern Copts in Egypt, it remains unclear whether selection occurred after their migration to Sudan or if it is also present in the Egyptian Coptic population. Also, the absence of population specific epidemiological data limits our ability to confirm whether the observed genetic adaptation directly correlates with malaria resistance in Copts. Future studies integrating serological testing of Duffy antigens with *P. vivax* malaria incidence data in both Coptic populations would help validate the hypothesized adaptive advantage.

Although the African continent harbors the greatest human genetic diversity, genomic studies remain biased toward European populations, with Africa still severely underrepresented in whole genome sequencing (WGS) datasets. This imbalance affects not only modern high-coverage genomes but is even more pronounced for ancient DNA, where data from Sudan and Egypt are nearly absent despite their key roles in human history. Such gaps limit our ability to detect deeper signals of population history and reduce the power to accurately estimate selection processes. While the number of available genomes in Africa is gradually increasing(20, 23, 77–80), Sudan and the broader Northern and Eastern African regions remain poorly covered. This study presents the first WGS dataset from Sudan and demonstrates the power of WGS to uncover the complex genetic landscape of a region of exceptional cultural and linguistic diversity. We show that historical admixture events have significantly shaped the genomes of its inhabitants, introducing genetic variation that, under local selective pressures, contributed to adaptation to malaria. These findings highlight the importance of including diverse and underrepresented populations in genomic research to fully capture human genetic variation and evolutionary history.

## Materials and methods

### Genome sequencing and variant filtering

DNA samples from five Sudanese populations were sequenced for whole genome data. In total, 125 samples from populations: Beja (N=25), Copts (N=25), Fulani (N=25), Fur (N=25) and Mahas (N=25) (*SI Appendix*, Dataset S1). All participants provided written informed consent, and the study was approved by the Ethical Committee of the University of Medical Sciences and Technology, Khartoum, Sudan.

The sequencing was performed with the Illumina HiseqX platform. Reads were aligned to the GRCh38 reference genome using BWA-MEM(81) with -Y -K 100000000. Duplicates were removed with bammarkduplicates from biobambam(82). The variant calling was done with HaplotypeCaller from GATK 4.1.9.0(83). We followed the GATK Best Practice Workflow for germline short variant discovery (SNPs + indels) from GATK/4.1.8.1(83) for the genotype calling and recalibration steps, ending with a total of 32,999,921 sites. The average coverage was 30.48X with a minimum coverage of 15X (*SI Appendix*, Dataset S1). For subsequent analysis, we kept 24,774,084 autosomal biallelic SNPs with read depth (DP) > 5, genotype quality (GQ) > 20 and missingness < 0.05. Those not present in dbSNPv156(39), gnomADv4.1(40) and the HGDP+1KG callset from gnomAD(41), were reported as novel and annotated with VEP(84). If the ancestral state was unavailable, the minor allele was assumed to be the derived allele. We removed samples with more than a first degree of relatedness using PLINKv1.9(85) and PRIMUSv1.9.0(86). A total of 117 out of 125 Sudanese samples passed the filter (*SI Appendix*, Dataset S1).

### Datasets preparation

For demographic analyses, we merged our dataset with high coverage African, Middle Eastern and European samples from the HGDP+1KG callset(41). In addition, we included other high coverage genomes from Northern Africa(20, 79), Nigeria(77), the Sahel(23) and mainland Africa(78) (*SI Appendix*, Dataset S2). The same filters for DP, GQ, relatedness and missingness described above were applied. Only overlapping variants between the datasets (5,814,532 SNPs) were kept for the *demography_dataset*.

For selection analyses, we merged our dataset with the HGDP+1KG, keeping the unrelated samples and all available QC-filtered variants, defining the latter as missing if not present in the other dataset (*selection_dataset*). Those with more than 5% of missingness were filtered out before phasing.

### Population structure and genetic differentiation

We computed the runs of homozygosity (ROHs) to estimate levels of inbreeding. For this we used PLINK with a minimum of 50 SNPs, a minimum length of 500 kb with a maximum gap of 100 kb, using a window size of 50 SNPs allowing maximum 5 missing calls and 1 heterozygous position in the scanning window. Additionally, ROHs were classified in length bins of 0-5-1 Mb, 1-2 Mb and >2 Mb, based on previous observations in the same ethnolinguistic groups and geographic region(19).

For Principal Component Analysis (PCA) and ADMIXTURE analyses in the *demography_dataset*, we filtered out with PLINK, variants with a Hardy-Weinberg exact test p-value >10^-7^, and those in linkage disequilibrium (LD) considering a maximum pairwise LD threshold (r^2^) of 0.5, and a window size of 200 SNPs shifting by 25 SNPs (1,427,188 SNPs). We performed PCA with smartpca from EIGENSOFTv6.0.1(87) and unsupervised ADMIXTUREv1.3.0(44) for 15 independent iterations with 2-15 ancestral components. The lowest cross-validation error of the ADMIXTURE analysis was for k=8. We used PONGv1.4.9(88) for visualization.

For a closer look at the genetic structure in Eastern Africa, we computed a PCA including genotype array data(18, 71, 89–91) for a total of 182,724 autosomal SNPs. To detect ancient genetic components, we ran ADMIXTURE for 15 iterations with 2-8 ancestral components, including samples of the AADRv54.1.p1(92) and ancient genomes from Northern Africa(93), for 123,477 autosomal SNPs.

### Haplotype estimation and admixture timing

To perform haplotype -based analyses we extracted from the *demography_dataset* the Affymetrix Axiom Genome-Wide Human Origins 1 Array SNPs (∼ 629,000)(94), to speed up the process, ending up with 407,502 SNPs. We phased this dataset with SHAPEITv4.2.2(95), using the haplotypes from the HGDP+1KG callset as reference panel, to get the maximum number of variants.

ChromoPainterv2(45) was used to reconstruct each sample as a mosaic of haplotypes from other samples. For this we estimated from chromosomes 1, 7, 14, and 20, the global mutation probability (M) (0.0037885098) and the switch rate (n) (464.8266192212) parameters.

Next, we used fineSTRUCTUREv2.1.0(45) on the chunkcounts ChromoPainter matrix to organize samples into genetic clusters, using 2 million MCMC iterations, 1 million burn-in iterations and sampling every 10,000 iterations. Three replicate dendrograms were generated using different seeds with these parameters. Genetic clusters were defined by evaluating the consistency of branching across the three replicates, assigning individuals based on the majority clustering when discrepancies occurred. All other settings were kept at their defaults (*SI Appendix*, Dataset S3). To quantify differentiation among these clusters, we computed the total variation distance (TVD) following the method in(96) (*SI Appendix*, Dataset S4). ChromoPainter was run again, setting which genetic clusters would perform as donors. The haplotype sharing proportions from different donor clusters were inferred for each recipient cluster using SOURCEFINDv2. We used 1 million MCMC iterations, 250,000 burn-in iterations and sampled every 7,500 iterations, averaging results from 100 independent runs (48).

The timing of admixture was estimated for the target genetic clusters with MALDERv1.0(47) and fastGLOBETROTTER(46). With the latter, we also estimated the source populations and their contributions to the admixture. DATESv4010(49) was used to infer the admixture time in Fulani between ancient populations from Western Africa (Cameroon_SMA)(50), Northern Africa (IAM)(51) and Eastern Africa (Kenya_LSA)(52, 53).

### Inference of local ancestry deviation

The *selection_dataset* was phased with SHAPEITv4.2.2(95), with haplotype reference data from the 1000 Genomes Project(97). Local ancestry inference was performed with RFMixv1.5.4(54), and genomic regions with local ancestry deviations (LAD) exceeding ±4.42 standard deviations (SD) from the genome-wide average, were identified as outliers(98). In some cases, a more relaxed threshold of ±2 SD was used for comparison. Fur (N=22) were used as a reference for Nilo-Saharan-like ancestry; Bedouin and CEU (N=164) for West Eurasian ancestry; and Yoruba (N=142) for Western African ancestry.

We extracted the genes within these regions from the UCSC Genome Browser on Human GRCh38/hg38(99). We reported the GO ontologies with FDR <0.05 for these genes with DAVID(100). Variants inside LAD regions were annotated with VEP(84). We defined as deleterious missense variants predicted to be probably or possibly damaging by PolyPhen-2(42), stop codon gain/loss, start codon loss, splice donor/acceptor, transcript ablation/amplification, feature elongation/truncation, frameshift variants and variants reported as pathogenic or likely pathogenic in ClinVar(43).

### Detection of positive selection signals

To detect signals of positive selection, we computed for the *selection_dataset* first the Hudson’s FSTs at a variant -level with PLINKv2.0(101). Next, we computed Population Branch Statistics (PBS)(64) for Beja, Coptic and Mahas populations, to find signals either associated with an African-like ancestry or specific for that population. For this purpose, we used Bedouins as the closest relatives from the Middle East and CEU as a European-like distant group. Significant peaks were defined inside the top 0.1% of PBS scores(102) with a minimum of 60 SNPs and a maximum distance of 20,000 bp between SNPs, to minimize the number of signals. We computed the rank p-values and extracted the most significant variant inside each peak. Genes within these peaks were extracted from the UCSC Genome Browser(99) and variants were annotated with VEP(84).

### Tracking evidence of local adaptation

We ran Relate(103) for all chromosomes on the phased *selection_dataset* to infer the genealogy of rs2814778 (T>C) considering 28 years per generation and 1.25×10^-8^ mutation rate. Coalescence rates of the Sudanese populations and the Effective Population Size (Ne) dynamics were also computed.

Next, we used CLUES2(65, 66) to infer the selection coefficient (s), its 95% confidence interval and the trajectory of the derived allele (C) for Copts, using 100 samples from Relate’s genealogies. Imputed phased chromosome 1 of ancient individuals from genetic clusters “Farmer_Iran” and “PostNeol_Levant_MedE” from Allentoft et al. (2024)(104) dataset, was used as point estimates for CLUES2’s hidden Markov model. We inferred selection between 0-45 generations ago, considering the average admixture event inferred in previous steps. We also applied a classical formula based on allele frequency change over time(67), assuming an additive effect of the Duffy-null allele(55, 68, 69). This allowed us to obtain an estimate of the selection coefficient without relying on the demographic assumptions required by CLUES2, providing a simpler point of comparison.

## Acknowledgements

This work was supported by grants PID2022-138755NB-I00 and PID2023-147621NB-I00 funded by MCIN/AEI/10.13039/501100011033 and “ERDF A way of making Europe”, by the European Union. We sincerely thank all participants who generously provided their DNA samples for this study, as well as Hanan Tahir and Abdulla Elkhawad of UMST for their assistance with Copts sample collection.

## Author contributions

L.V. -V., D.C. and A.M.A. designed the study. L.V.-V. conducted the analysis. L.V.-V., J.G.-C.,

D.C. and A.M.A interpreted the data. L.V.-V. prepared the manuscript with help of D.C.; and L.V. -V., J.G.-C., J.P.-M., E.B., A.M.A., M.G.N., D.C. and H.Y.H. contributed with the sampling and discussing the results. All authors revised and approved the manuscript.

## Ethics declarations

### Competing interests

The authors declare no competing interests.

## Data availability

WGS data from Sudan is available at the European Genome-phenome Archive (EGA), under accession number EGAD00001015636.

## Notes

### Competing Interest Statement

The authors have declared no competing interest.

### Funding Statement

This study was funded by grants PID2022-138755NB-I00 and PID2023-147621NB-I00 funded by MCIN/AEI/10.13039/501100011033 and ERDF A way of making Europe, by the European Union.

### Author Declarations

The Ethical Committee of the University of Medical Sciences and Technology, Khartoum, Sudan, gave ethical approval for this work.

