## Supplementary material for "Sudan’s complex genetic admixture history drives adaptation to malaria in Sudanese Copts": SI_Appendix.pdf

### Summary

**Figures (S1 to S9)** **3**
**Tables (S1 to S2)** **12**
**Datasets (S1 to S6)** **14**

#### Figures (S1 to S9)

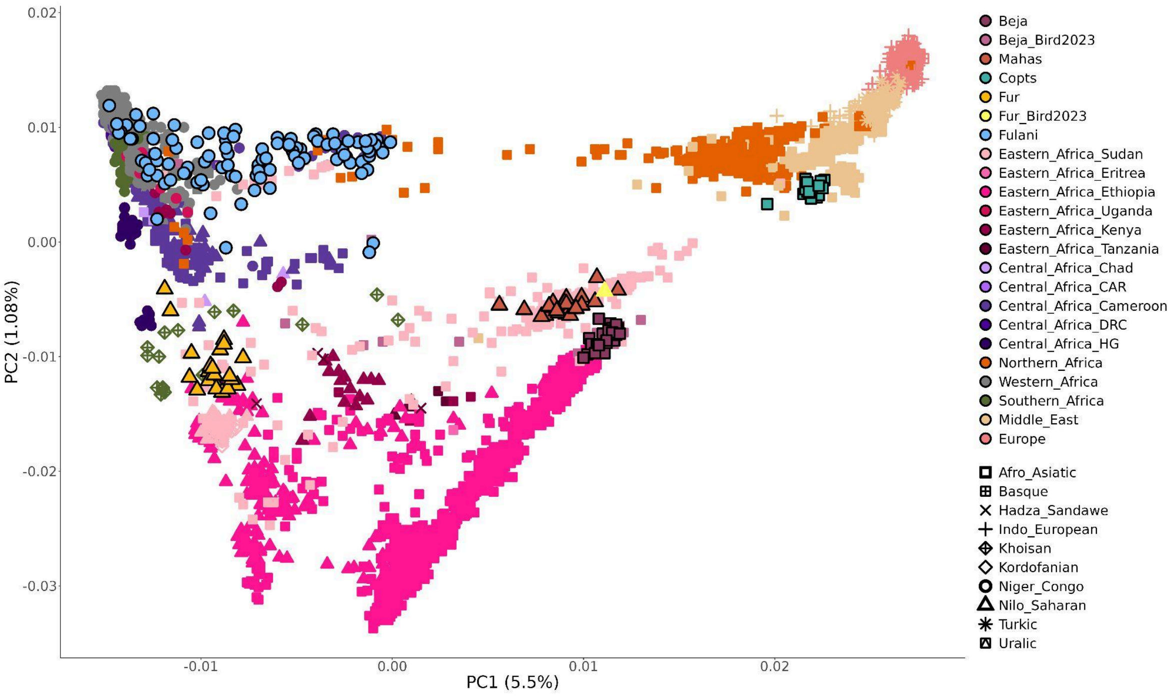


**Fig. S1** Genetic and linguistic structure in East Africa. Principal Component Analysis (PCA) computed for a dataset including genotype array data of many Eastern African and neighboring populations. Samples are colored by population or country. Shapes define language families.

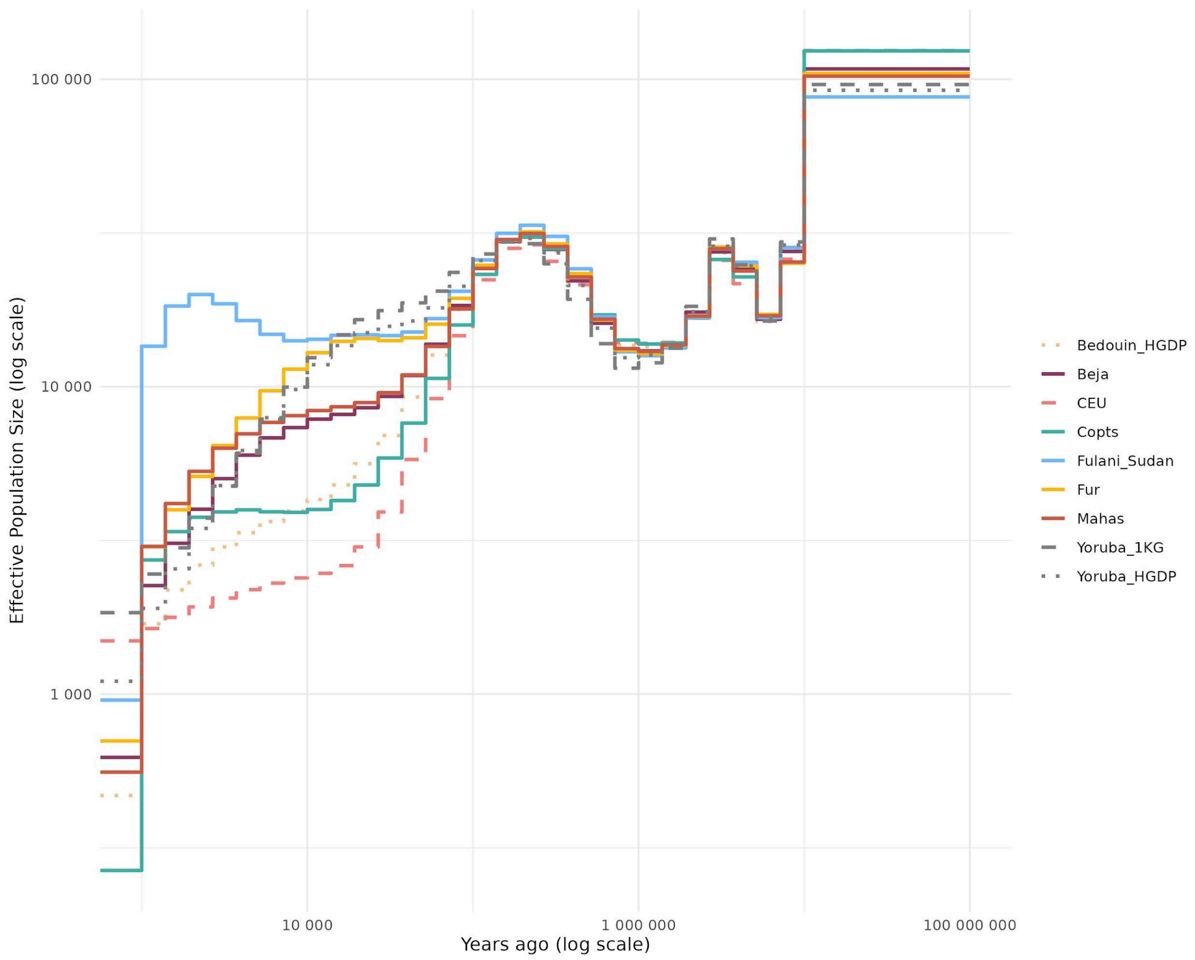


**Fig. S2** Effective population size dynamics. Effective population size (Ne) through time obtained from the coalescence rates computed with Relate assuming a mutation rate of 1.25 × $10^{-8}$ and 28 years/generation. Axes are in log scale for better visualization.

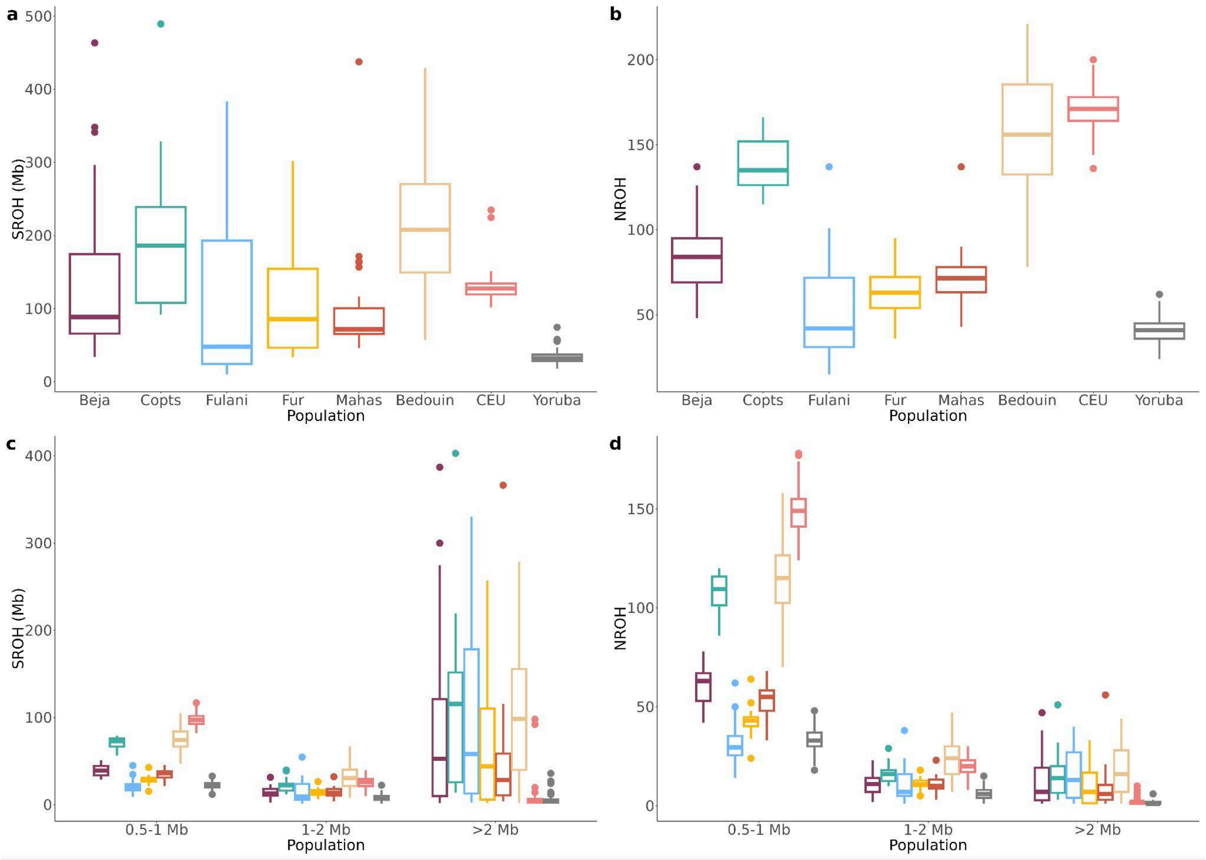


**Fig. S3** Runs of homozygosity (ROH) of at least 500kb. **a)** total sum of ROHs (SROH) by population. **b)** total number of ROH (NROH) by population. **c)** SROH by different length category and population. **d)** NROH by different length category and population.

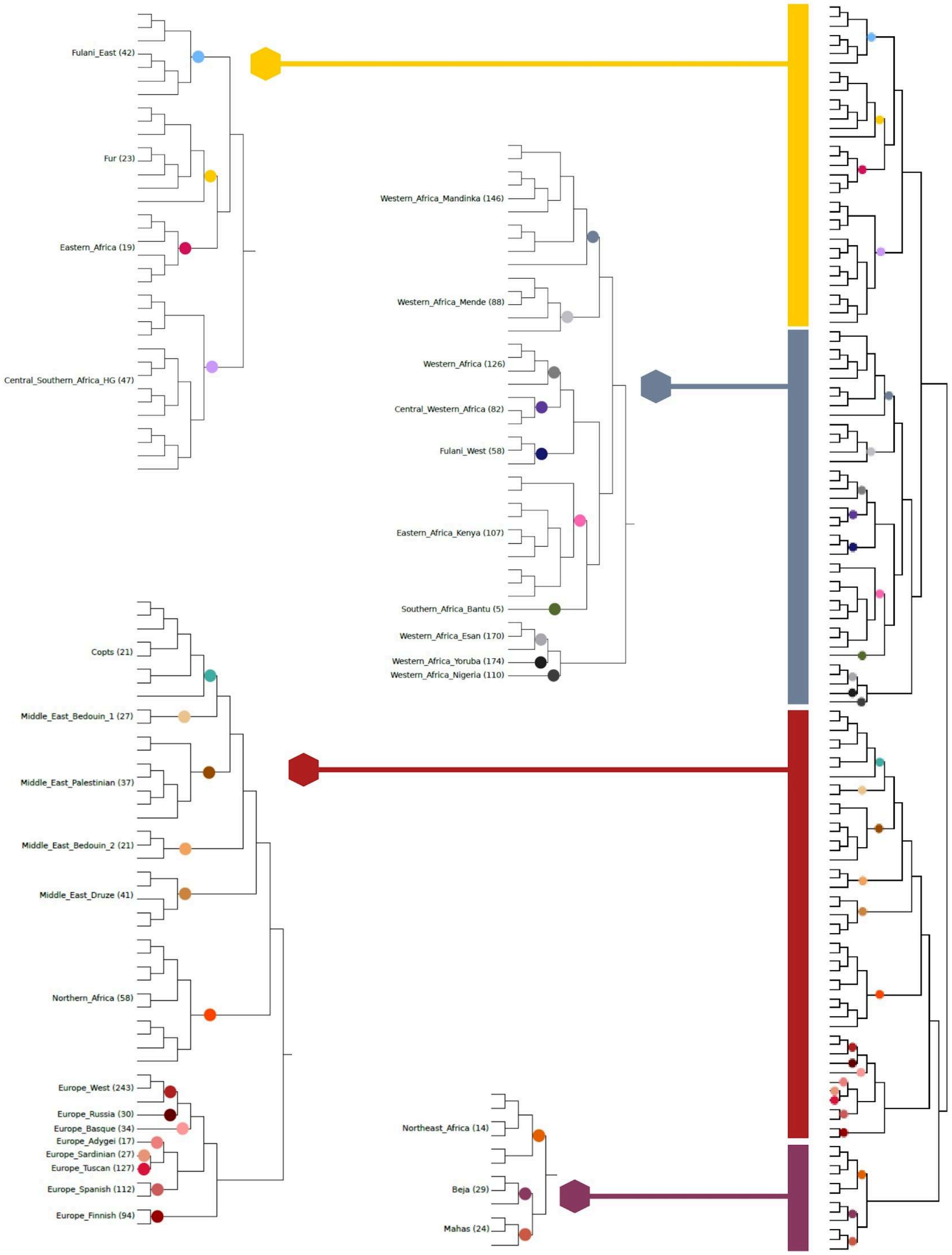


**Fig. S4** FineSTRUCTURE dendrogram. Dendrogram computed with fineSTRUCTURE using the chunkcounts coancestry matrix from ChromoPainter. It was used to group the samples based on genetic similarity. Four general branches are represented from top to bottom: Eastern Africa, Western/Central/Southern Africa, Europe/Middle East/Northern Africa and Northern/Eastern Africa. A closer look shows 31 genetic clusters defined at the height of each dot.

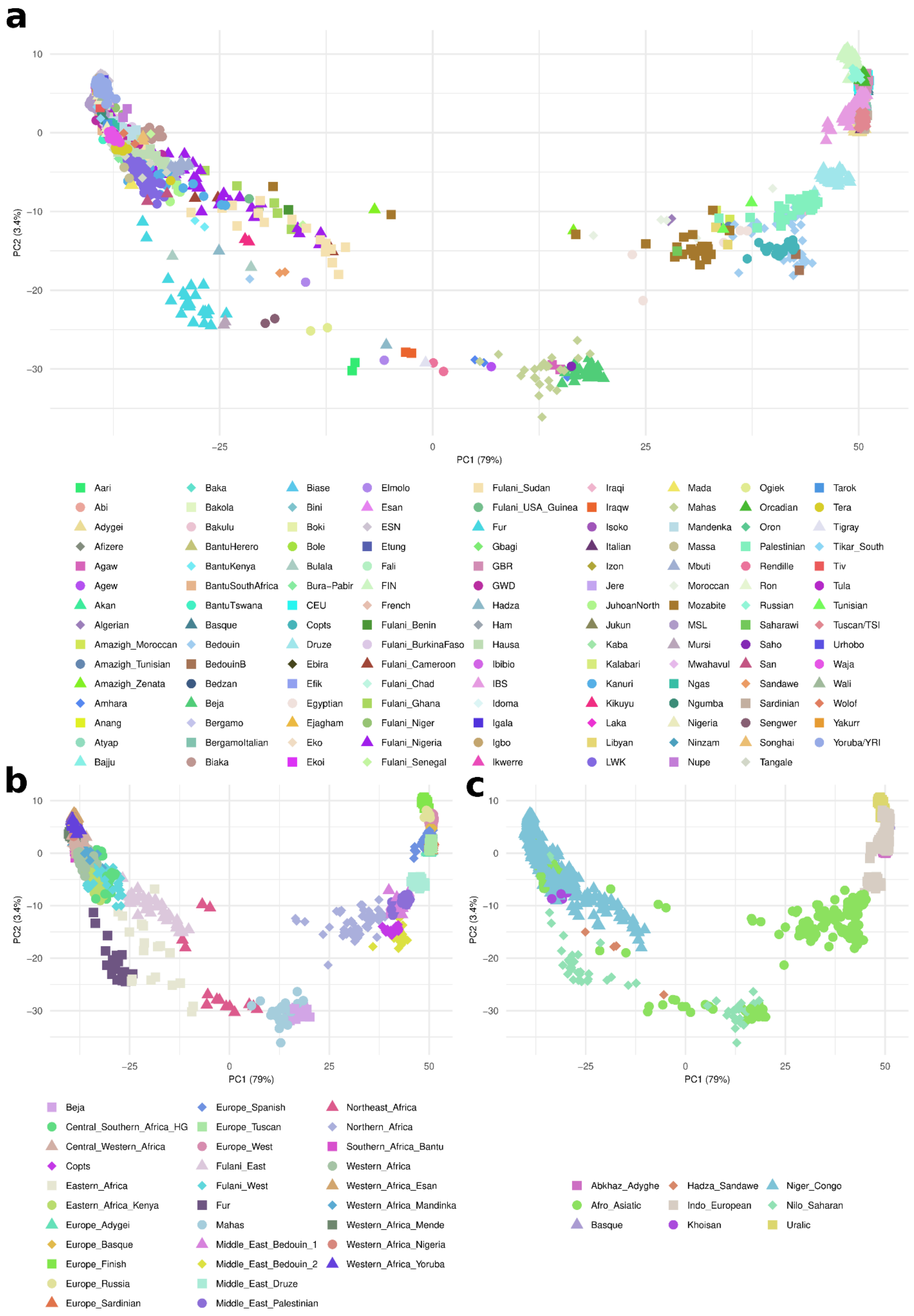


**Fig. S5** PCAs computed for the chunkcounts output matrix from ChromoPainter. **a)** Individuals classified by population labels. **b)** Classified by genetic clusters. **c)** Classified by language family

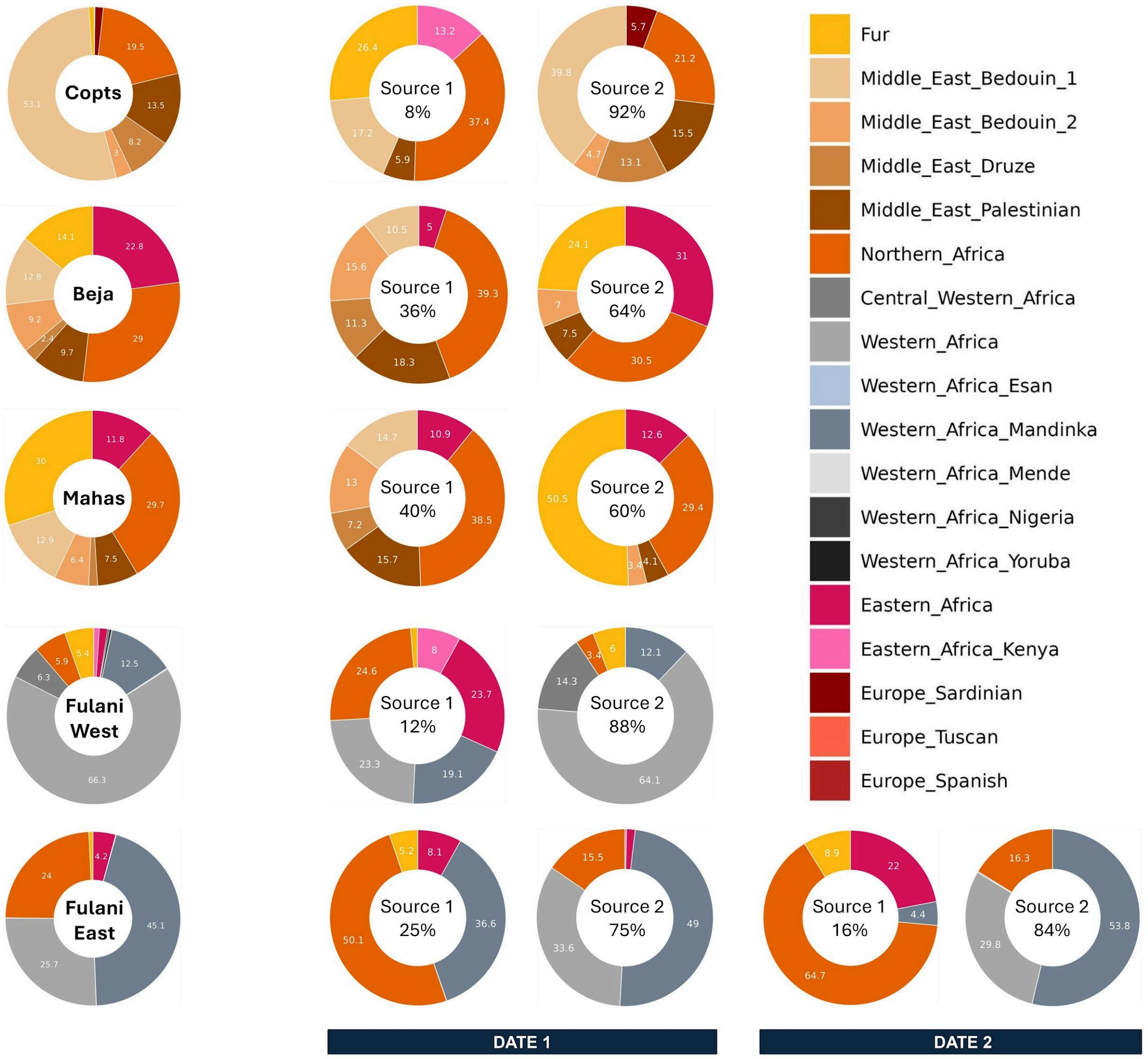


**Fig. S6** Ancestry proportions of the target and source populations. On the left, the target (recipient) populations with their ancestry composition calculated with SOURCEFINDv2. On the right, the ancestry proportions and the contributions of the source populations participating in the admixture, computed with fastGLOBETROTTER. Percentages smaller than 2% are not shown.

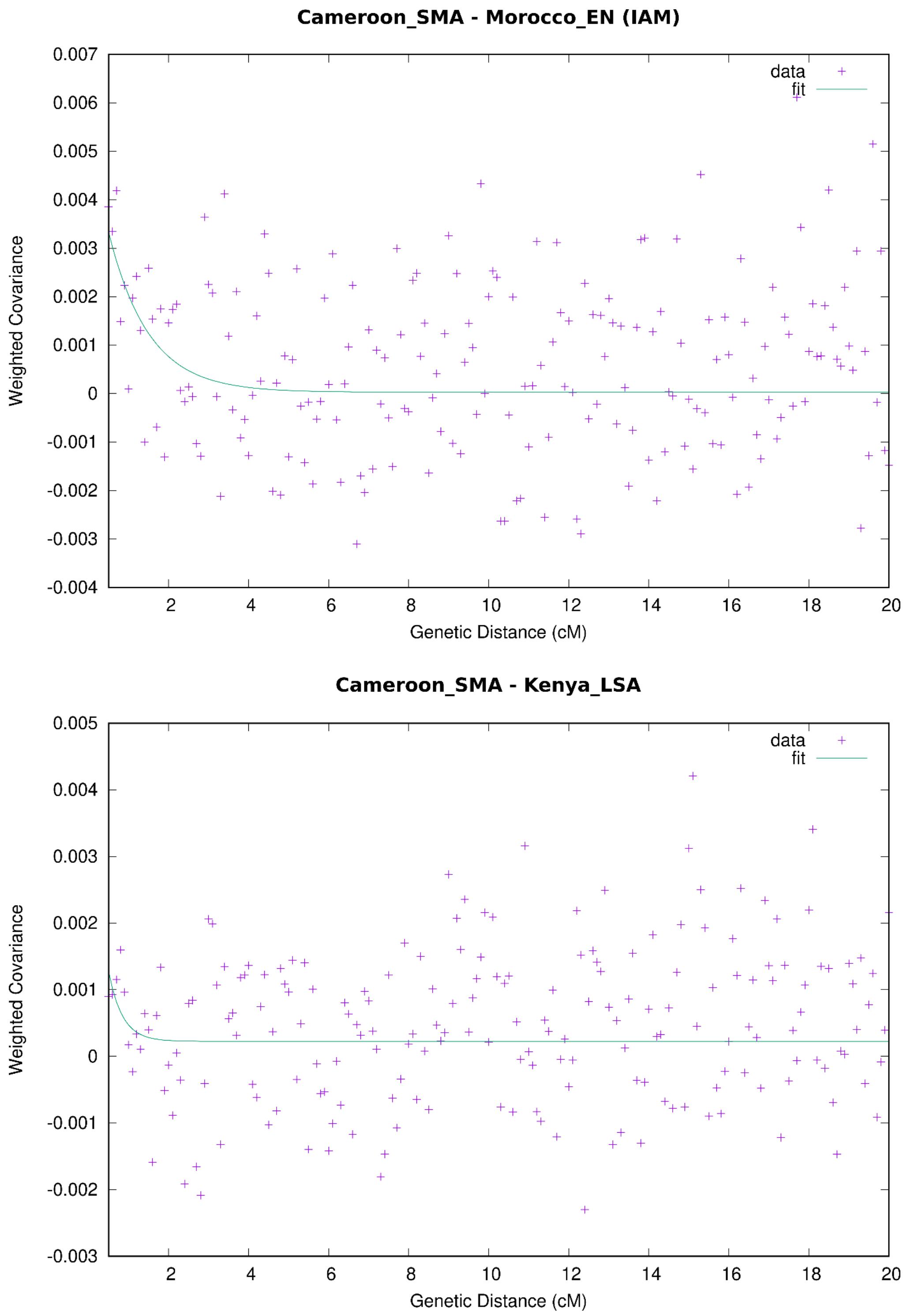


**Fig. S7** Ancestry covariance decay curves from DATESv4010 estimates. Top: admixture test between ancient Cameroon individuals and Moroccan Early Neolithic (IAM). Bottom: admixture test between ancient Cameroon and Later Stone Age individuals from Kenya.

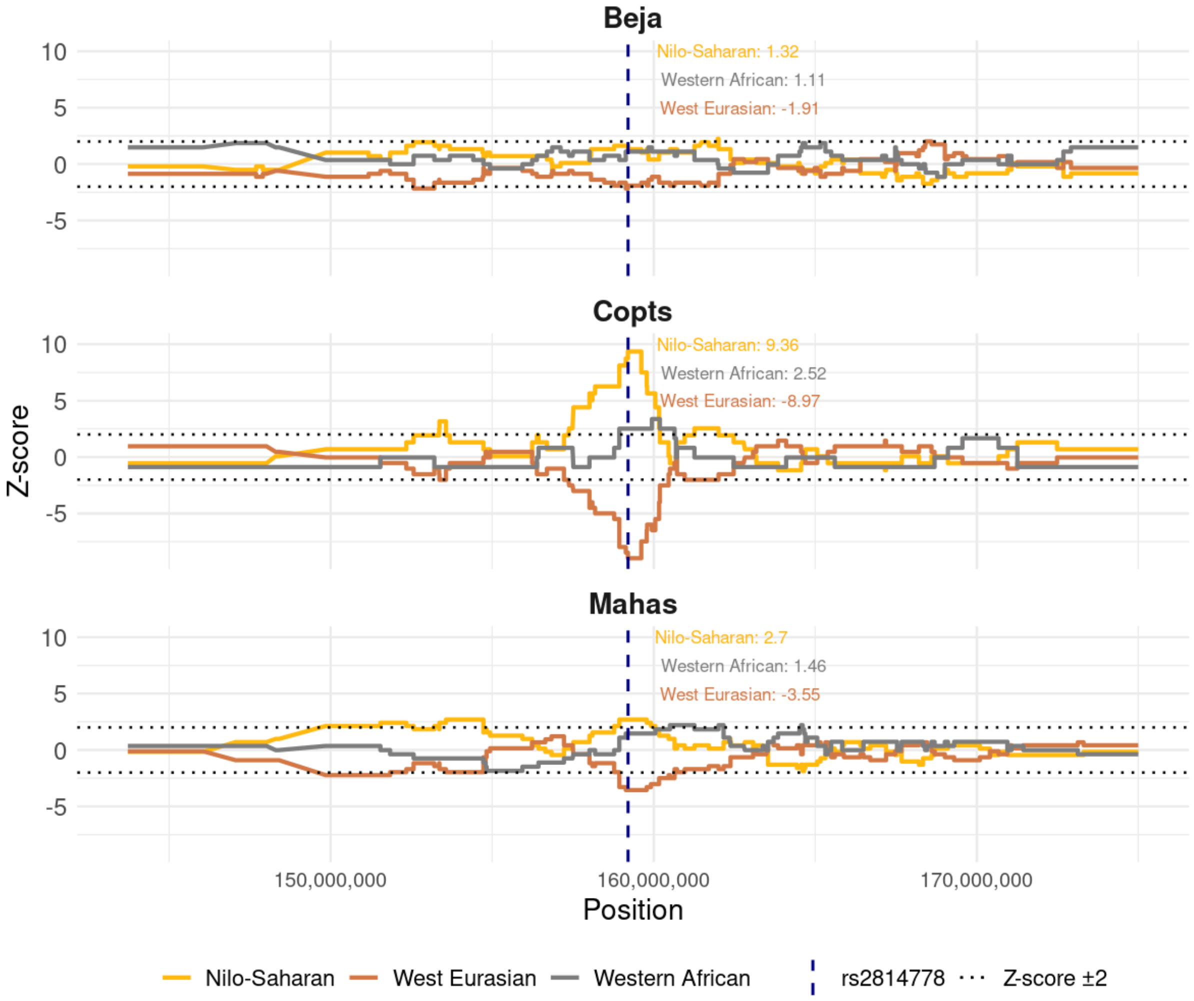


**Fig. S8** Local ancestry inference around the Duffy-null variant. Region of chromosome 1 containing the selection signal observed in Copts (Fig. 3), shown here for Beja, Copts and Mahas, using a z-score threshold of ± 2. The z-scores in the plots correspond to the Duffy-null variant for each ancestry.

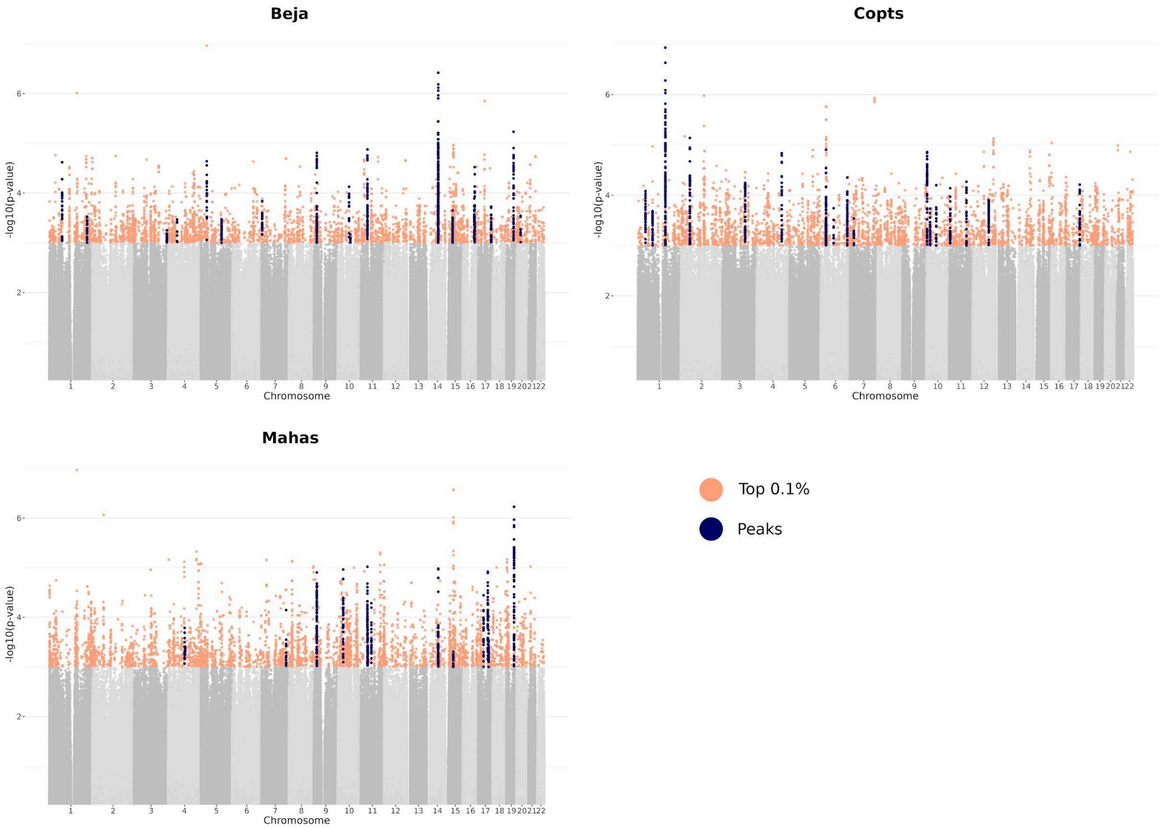


**Fig. S9** Manhattan plots for the PBS analysis. In orange SNPs with a PBS value in the top 0.1%. In blue, significant peaks, defined considering a minimum of 60 SNPs in the top 0.1% of PBS scores with a maximum distance of 20,000 bp between SNPs.

#### Tables (S1 to S2)

**Table S1.** Unreported SNPs. This table includes the number of unreported variants in total and by population, which of them are deleterious and their prediction, which have a derived allele frequency > 0.05 and in which genes are found.

| **population** | **unreported** | **deleterious** | **counts** | **highlights (derived freq > 0.05)** | **gene_highlights** |
|---|---|---|---|---|---|
| Fur | 246480 | 419 | missense_variant + PolyPhen=possibly_damaging: 167; missense_variant + PolyPhen=probably_damaging: 203; stop_gained: 27; splice_donor_variant: 8; splice_acceptor_variant: 7; stop_lost: 2; start_lost: 5 | chr3:53876707 (Fur:0.07) stop_gained | ACTR8 |
| Fulani | 146566 | 278 | missense_variant + PolyPhen=possibly_damaging: 110; splice_acceptor_variant: 5; missense_variant + PolyPhen=probably_damaging: 140; stop_gained: 15; splice_donor_variant: 8 | chr7:6410271 (Fulani:0.06 missense_variant + PolyPhen=probably_damaging(0.995) | DAGLB |
| Copts | 154552 | 207 | missense_variant + PolyPhen=probably_damaging: 101; missense_variant + PolyPhen=possibly_damaging: 80; splice_donor_variant: 6; stop_gained: 13; splice_acceptor_variant: 7 | chr1:117018165 (Copts:0.07) missense_variant + PolyPhen=probably_damaging(0.982)<br>chr6:63713538 (Copts:0.07) stop_gained<br>chr9:18777201 (Copts:0.07) missense_variant + PolyPhen=possibly_damaging(0.835) | CD101<br>PHF3<br>ADAMTSL1 |
| Mahas | 215557 | 343 | missense_variant + PolyPhen=probably_damaging: 184; missense_variant + PolyPhen=possibly_damaging: 104; splice_acceptor_variant: 12; stop_gained: 31; splice_donor_variant: 10; start_lost: 1; stop_lost: 1 | chr4:72295684 (Mahas:0.08) missense_variant + PolyPhen=possibly_damaging(0.45)<br>chr12:43446651 (Mahas:0.06) missense_variant + PolyPhen=probably_damaging(0.994); ClinVar=2269716<br>chr12:70609315 (Mahas:0.06) missense_variant + PolyPhen=probably_damaging(0.917) | ADAMTS3<br>ADAMTS20<br>PTPRB |
| Beja | 216550 | 278 | missense_variant + PolyPhen=probably_damaging: 148; stop_gained: 12; missense_variant + PolyPhen=possibly_damaging: 99; splice_donor_variant: 9; splice_acceptor_variant: 7; other + ClinVar_CLNSIG=Likely_pathogenic: 1; start_lost: 1; stop_lost: 1 | chr7:98240879 (Beja:0.06) missense_variant + PolyPhen=possibly_damaging(0.681)<br>chr13:27435770 (Beja:0.06) stop_gained<br>chr16:30655933 (Beja:0.06) missense_variant + PolyPhen=possibly_damaging(0.707)<br>chr19:55810101 (Beja:0.06) missense_variant + PolyPhen=probably_damaging(1) | TECPR1<br>GTF3A+MTIF3<br>PRR14<br>NLRP11 |
| Shared between 2 populations | 53152 | 13 | missense_variant + PolyPhen=possibly_damaging: 6; missense_variant + PolyPhen=probably_damaging: 6; splice_donor_variant: 1 | chr14:64541996 (Mahas:0.02, Copts:0.07) missense_variant + PolyPhen=probably_damaging(0.94) | HSPA2 |
| Shared between 3 population | 30154 | 4 | missense_variant + PolyPhen=possibly_damaging: 4 | chr17:17164359 (Mahas:0.06,Fur:0.02,Copts:0.02) missense_variant + PolyPhen=possibly_damaging(0.815) | MPRIP |
| Shared between 4 population | 24537 | 0 | | | |
| Shared between 5 population | 49062 | 0 | | | |
| **Total** | **1136610** | **1542** | missense_variant + PolyPhen=possibly_damaging: 570; missense_variant + PolyPhen=probably_damaging: 782; splice_acceptor_variant: 38; stop_gained: 98; splice_donor_variant: 42; other + ClinVar_CLNSIG=Likely_pathogenic: 1; stop_lost: 4; start_lost: 7 | | |

**Table S2.** Dates of admixture. Average admixture dates and their 95% confidence intervals computed with fasGLOBETROTTER and MALDER. With fasGLOBETROTTER we inferred more than one event of admixture (date) for the *Fulani_East* cluster. The last column is the average between methods.

| test_cluster | date | method | mean | lower_ci | upper_ci | average_methods |
|---|---|---|---|---|---|---|
| Beja | 1 | MALDER | 58,403 | 57,513 | 59,292 | 54,031 |
| | 1 | fastGLOBETROTTER | 49,659 | 48,938 | 50,381 | |
| Copts | 1 | MALDER | 51,796 | 50,402 | 53,191 | 44,906 |
| | 1 | fastGLOBETROTTER | 38,016 | 36,875 | 39,158 | |
| Fulani_East | 1 | MALDER | 16,806 | 15,784 | 17,829 | 15,285 |
| | 1 | fastGLOBETROTTER | 13,763 | 12,889 | 14,637 | |
| | 2 | fastGLOBETROTTER | 80,070 | 74,925 | 85,215 | - |
| Fulani_West | 1 | MALDER | 8,125 | 7,440 | 8,810 | 15,641 |
| | 1 | fastGLOBETROTTER | 23,157 | 22,734 | 23,580 | |
| Mahas | 1 | MALDER | 26,629 | 25,779 | 27,478 | 30,185 |
| | 1 | fastGLOBETROTTER | 33,741 | 33,272 | 34,210 | |

#### Datasets (S1 to S6)

**Dataset S1.** Sample information. This table includes all the samples from the dataset (N=125) and their information. (1) Samples ID, (2) Population, (3) Population Super/Sub-group, (4) Language Family, (5) Language Subfamily, (6) Socio-economical Activities (7) Coordinates, (8) City/Sampling location, (9) Average Coverage, (10) if they were removed for more than first degree of relatedness.

**Dataset S2.** Reference panel. This table includes all the populations or ethnolinguistic groups used for the main analyses with WGS data and their information. (1) Population, (2) Sample Size (N), (3) Country, (4) Language Family, (5) Reference.

**Dataset S3.** Genetic clusters defined with fineSTRUCTURE. This table includes all the genetic clusters defined with fineSTRUCTURE with column (1) cluster name, (2) if it is used as donor or only as recipient in ChromoPainter, (3) original population names, (4) number of individuals from each population in column 3 included in that cluster and (5) size of the genetic cluster.

**Dataset S4.** Total variation distance (TVD) between the 31 genetic clusters defined with fineSTRUCTURE.

**Dataset S5.** Regions with local ancestry deviation. This table contains all the regions with local ancestry deviation (LAD) (|z-score| ≥ 4.42). (1) Population, (2) chromosome, (3) start position, (4) end position, (5) ID region in LAD, (6) ancestry in LAD, (7) average ancestry proportion of the region, (8) average ancestry proportion of the genome, (9) average z-score, (10) chromosomal location, including whether the variant is situated at the telomeres (start or end) or near the centromere, (11) coding genes englobed, (12) number of variants inside the region, (13) number of deleterious variants, (14) annotation of the deleterious variants, (15) gene ontologies with a p-value after FDR correction <0.05. Rows in red indicate peaks that were discarded for being prone to technical artifacts.

**Dataset S6.** PBS peaks. This table contains all the peaks after computing Population Branch Statistics (PBS), considering a minimum of 60 SNPs in the top 0.1% of PBS scores with a maximum distance of 20,000 bp between SNPs. (1) Population, (2) chromosome, (3) start position, (4) end position, (5) ID peak, (6) location in the chromosome, (7) coding genes englobed, (8) number of variants inside the peak, (9) variant with the highest value inside the peak, (10) PBS value of the top variant, (11) p-value of the top variant, (12) annotation of the top variant, (13) number of deleterious variants, (14) annotation of the deleterious variants.
